# Monitoring for COVID-19 by universal testing in a homeless shelter in Germany: a prospective feasibility cohort study

**DOI:** 10.1101/2020.10.04.20205401

**Authors:** Andreas K. Lindner, Navina Sarma, Luise Marie Rust, Theresa Hellmund, Svetlana Krasovski-Nikiforovs, Mia Wintel, Sarah M. Klaes, Merle Hoerig, Sophia Monert, Rolf Schwarzer, Anke Edelmann, Gabriela Equihua Martinez, Frank P. Mockenhaupt, Tobias Kurth, Joachim Seybold

## Abstract

**Background:** Living conditions in homeless shelters may facilitate the transmission of COVID-19. Social determinants and pre-existing health conditions place homeless people at increased risk of severe disease. Described outbreaks in homeless shelters resulted in high proportions of infected residents and staff members. In addition to other infection prevention strategies, regular shelter-wide (universal) testing for COVID-19 may be valuable, depending on the level of community transmission and when resources permit.

**Methods:** This was a prospective feasibility cohort study to evaluate universal testing for COVID-19 at a homeless shelter with 106 beds in Berlin, Germany. Co-researchers were recruited from the shelter staff. A PCR analysis of saliva or self-collected nasal/oral swab was performed weekly over a period of 3 weeks in July 2020. Acceptability and implementation barriers were analyzed by process evaluation using mixed methods including evaluation sheets, focus group discussion and a structured questionnaire.

**Results:** Ninety-three out of 124 (75%) residents were approached to participate in the study. Fifty-one out of the 93 residents (54.8%) gave written informed consent. High retention rates (88.9% – 93.6%) of a weekly respiratory specimen were reached, but repeated collection attempts, as well as assistance were required. A self-collected nasal/oral swab was considered easier and more hygienic to collect than a saliva specimen. No resident was tested positive. Language barriers were the main reason for non-participation. Flexibility of sample collection schedules, the use of video and audio materials, and concise written information were the main recommendations of the co-researchers for future implementation.

**Conclusion:** Voluntary universal testing for COVID-19 is feasible in homeless shelters. Universal testing of high-risk facilities will require flexible approaches, considering the level of the community transmission, the available resources, and the local recommendations. Lack of human resources and laboratory capacity may be a major barrier for implementation of universal testing, requiring adapted approaches compared to standard individual testing. Assisted self-collection of specimens and barrier free communication may facilitate implementation in homeless shelters. Program planning must consider homeless people’s needs and life situation, and guarantee confidentiality and autonomy.

## Background

People experiencing homelessness represent a vulnerable group with complex needs. Due to poor linkage-to-healthcare as well as lack of fulfilment of basic needs, they have higher occurrence of chronic mental and physical conditions, and higher mortality rates [1-3]. Exposure to severe acute respiratory syndrome coronavirus type 2 (SARS-CoV-2) might negatively affect their health, and further magnify these social and health inequalities [4]. Social determinants and pre-existing health conditions place homeless people at higher risk of severe coronavirus disease 2019 (COVID-19) [5, 6]. The mobile nature of the community, high rates of substance abuse, informal sector employment or fear of involuntary hospitalization should be considered for screening, infection prevention, quarantining and treatment [4, 7, 8]. Access to health information, compliance with distance and hygiene rules, or self-isolation in case of symptoms can be a challenge for homeless people [9].

Congregate living settings in community shelters for homeless people that have shared bedrooms and sanitary facilities, could facilitate the transmission of COVID-19. The German notification system does not allow identification of homeless status and to our knowledge there is no data on COVID-19 among homeless people in Germany. In Canada, several outbreaks in homeless shelters were observed [10]. In the USA, SARS-CoV-2 PCR testing of an adult homeless shelter population shortly after the identification of a COVID-19 cluster in that facility yielded an alarming 36% of positive tests [11]. At the time of identification, the vast majority of new cases was asymptomatic. COVID-19 outbreaks were also detected at other homeless shelters in the USA, with high proportions of infected residents and staff members when testing followed identification of a cluster [7, 12, 13]. Universal testing for COVID-19 at shelters is considered valuable when clusters occur. Moreover, preemptive testing in shelters can be considered, especially when transmission is increasing in the general population [12, 14, 15].

COVID-19-related lockdown measures, contact restrictions and a decline of volunteer staff in homeless support services contributed to a general decrease of support of the homeless population globally [16, 17]. Provision of necessary measures for homeless people during the pandemic, e.g., shelter, basic needs and health care, is being demanded by the United Nations Human Rights Commission and other organizations [18-22]. The German federal working group for homeless assistance explicitly called for the initiation of continuously open (24/7) shelters for homeless people [23]. To address these issues, the Senate of Berlin has opened three timely limited 24/7 shelters for people experiencing homelessness in May 2020, as well as a quarantine unit for SARS-CoV2 infected individuals [24].

Reverse transcription-polymerase chain reaction (RT-PCR) testing for SARS-CoV-2 is the current gold standard [25]. Oropharyngeal or nasopharyngeal swabs for specimen collection are frequently perceived as uncomfortable and sometimes painful by the tested individuals. Compliance with repeated testing, applying oropharyngeal or nasopharyngeal swabs is likely to be difficult. Moreover, it requires numerous resources, like qualified staff and personal protective equipment. Evidence on the validity of less invasive sampling methods such as saliva collection or swabs taken from the nasal mid-turbinate or anterior nares, is increasing and was taken into account for example by the European Centre for Disease Prevention and Control, and the US Centers for Disease Control and Prevention [26, 27]. These methods can also be performed by individuals themselves.

The objectives of this study were 1) to assess the feasibility of monitoring for COVID-19 by universal testing in a homeless shelter and 2) to assess the feasibility of the study methods, especially in regard to the specimen collection and work load.

The results should serve as a basis for future monitoring concepts, e.g., for the reopening of emergency overnight shelters for homeless people in Berlin during the winter season 2020/21 (in the previous winter season > 40 shelters with a capacity of > 1200 beds [28]).

## Methods

### Design, setting and participants

This was a prospective, feasibility cohort study with a mixed methods approach. Homeless people were recruited in one of three temporarily established 24/7 shelters in Berlin, Germany. The aim of the collaborative project between the Charité – Universitätsmedizin Berlin and the operator of the shelter, Berliner Stadtmission, was to enable shelter staff to conduct the study with a high degree of ownership and to integrate the study in their routine activities. The 24/7 shelter has 106 beds with shared bedrooms for up to 6 homeless adults. There is a fluctuation of shelter residents with about 10 new admissions per week. To be hosted, residents must register on-site and fulfill requirements during their stay, such as mandatory temperature measuring and compliance with certain schedules. On the same location, there is an associated healthcare center with a newly established COVID-19 quarantine unit with 16 beds, for individuals not requiring hospitalization. The study was conducted over a period of 3 weeks between 9 July and 29 July 2020.

### Co-researcher team and recruitment

The study was initiated and supervised by a team of infectious diseases and public health professionals. The implementation was carried out by shelter and quarantine unit staff (co-researchers). The multilingual co-researcher team (German, English, French, Spanish, Russian, Polish, Romanian and Lithuanian) consisted of two coordinators, social work assistants, nurses, medical students, and physicians. All co-researchers were trained with a focus on good clinical practice (GCP) standards considering the vulnerability of the shelter residents. As part of the process evaluation, daily meetings during the first week enabled timely decision on readjustments of the monitoring design. The co-researchers also initiated an online communication platform to disseminate instructions and advice from the coordinators, to provide daily updates, clarify questions and share experiences.

Oral and written study information was provided to the residents to obtain written consent. Potential participants were informed in their native languages if available. Some residents were informed in groups (minimum of 2 persons), with the possibility of a personalized explanation afterwards. The consent form was available in German, English, French, Russian, Polish, Bulgarian, Romanian and Arabic.

All shelter residents, independently of symptoms, were eligible for the study. Residents were excluded if the communication was not sufficient to obtain informed consent to participate. Shelter staff in direct contact with the residents was also offered participation in this study, but their results were not included in the analysis.

### Specimen collection and analysis

The monitoring concept aimed to obtain a self-collected respiratory specimen of each resident on a weekly basis irrespective of symptoms. During the first week, saliva was used as a specimen. Residents were asked during the weekday’s morning round (6:30 am to 9 am) to spit into a tube through a straw. The procedure was guided by an instruction leaflet provided [Additional file 1], with additional staff support if needed. The targeted volume of saliva, marked on the tube, was 2 ml. The specimens were transported to the laboratory within 3-6 hours of collection.

During the second and third week, a self-collected swab of wiping tongue, buccal mucosa and anterior nares was used, guided by an instruction leaflet [Additional file 2], with additional staff assistance if needed. The specimen was taken with the eSwab (Copan Diagnostics, Inc., USA) system with a nylon-flocked swab and liquid modified Amies medium. The study protocol permitted a collection regardless of the time of the day, as the specimen was in a tube with media and a short pre-analytical time was not of concern.

The participants were informed that in case of symptoms suspicious for COVID-19 the self-collected specimen did not replace a medical consultation at the ambulatory clinic and a swab taken by a health professional.

All samples were visually inspected to assess the proper closure of the tube, apparent abnormalities of the sample, and to perform sample volume estimation.

### RT-PCR analysis method

Especially saliva samples arrived in highly viscous condition in the laboratory. Pretreatment of these samples with DTT (Dithiothreitol) was carried out before RNA extraction on MagNA Pure 96 followed by real-time reverse transcriptase PCR (RT-PCR) on LightCycler 480 targeting the E-gene (LightMix^®^ SarbecoV E-gene kit, Tib molbiol). eSwab samples were analyzed after addition of 1 ml Roche cobas PCR Medium on cobas 6800/8800 using the CE labeled cobas^®^ SARS-CoV-2 assay (Roche) according to the manufacturer’s guidelines. An internal control in each sample as well as positive and negative controls were included in every run of both assays.

### Outcomes and measurements

Table 1 provides a summary of outcomes, measures/approaches, and methods of analysis corresponding to each objective. Regarding residents’ acceptability, the main measures were recruitment and retention rates. Recruitment rate was defined as the number of shelter residents that consented per number of residents who could be approached for participation. Retention rate was defined as the number of residents who were monitored with analysis of a respiratory specimens on a weekly basis compared with the number of recruited residents that were still living in the shelter during that week.

**Table 1.** Summary of outcomes, measures/approaches, and methods of analysis corresponding to each study objective.

| **Objectives** | **Outcomes** | **Measures/approaches** | **Methods of analysis** |
| --- | --- | --- | --- |
| Feasibility of study implementation | Residents' acceptability | - Recruitment rate<br>- Retention rate<br>- Evaluation forms<br>- Focus group (staff) | - Descriptive analysis<br>- Content analysis |
|  | Implementation barriers and facilitators | - Evaluation forms<br>- Focus group (staff)<br>- Continuous feedback *via* online communication platform, calls, and field visits | - Descriptive statistics<br>- Content analysis |
|  | Staff acceptability | - Evaluation form<br>- Focus group (staff) | - Descriptive statistics<br>- Content analysis |
| Feasibility of study methods | Specimen acceptability | - Visual inspection of specimen<br>- Evaluation forms<br>- Focus group (staff) | - Descriptive statistics<br>- Content analysis |
|  | Workload | - Evaluation form<br>- Focus group (staff) | - Descriptive statistics<br>- Content analysis |

Information on implementation barriers was collected with two evaluation forms that were developed together with the co-researchers. The forms were filled out by the co-researchers directly after the informed consent interview and specimen collection. The main variables recorded for the informed consent process were: language, duration of the interview, questions and doubts mentioned by the potential participant, difficulties as perceived by the co-researcher team, and suspected reasons for non-participation. The main variables recorded for the specimen collection were: number of attempts needed to collect the sample, difficulties reported by the residents, difficulties observed by the co-researcher team, and reason for non-collection of a specimen.

A final focus group with the co-researchers took place to discuss the feasibility of the overall approach. Based on experiences from the feasibility study, it was discussed to which extend the monitoring design needs to be refined for future implementation in similar settings. A quantitative structured questionnaire addressed recommendations for further implementation, team composition, sample collection, the essential content of the informed consent interview, assumed challenges for further implementation and critical appraisal of the pilot study. Moreover, protocols and notes of all meetings and trainings, the focus group discussion, and the online communication platform of the co-researcher team were considered for process evaluation and continuous adaption of the study design.

### Data analysis

We used descriptive statistics to summarize recruitment, retention, and baseline characteristics, as well as to compare residents that consented and declined participation by age, sex and language of the consultation. The data analysis of the process evaluation is based on the framework method [29]. It is applicable for multi-disciplinary health research teams that include practical involvement as well as leadership from experienced qualitative researchers. Co-researchers were involved in the analysis process. After a first assessment of the qualitative data, categories and descriptive labels (codes) were applied to the data [see Additional file 3]. A combined approach (deductive and inductive) was used for anticipating unexpected aspects or events. Framework analysis includes an ongoing interplay between data collection, analysis, and theory development. Further information, according to the consolidated criteria for reporting qualitative research (COREQ), is given in Additional file 4.

### Ethics

This study was approved by the ethics committee of the Charité – Universitätsmedizin (No.: EA4/141/20). The co-researcher team was sensitized to the dependencies between the co-researchers and the shelter residents that might cause research participation coercion. Residents were explicitly informed that positive RT-PCR test results for SARS-CoV-2 would have isolation as a possible consequence and would be immediately reported to the responsible local health authorities according to the infectious disease act. Notification includes personal details such as name and birth date of the person.

## Results

### Recruitment and barriers

Due to fluctuation, 124 residents were living in the shelter with 106 beds during the three-week study period. Fig. 1 shows the study flow with reasons of non-recruiting and non-retaining of shelter residents for a weekly respiratory specimen. Ninety-three out of 124 (75%) residents were approached to participate in the study. Thirty-one out of 124 (25%) could not be approached, either because they were not available during recruiting times, or because language mediation was needed and was unavailable.

**Figure 1.**
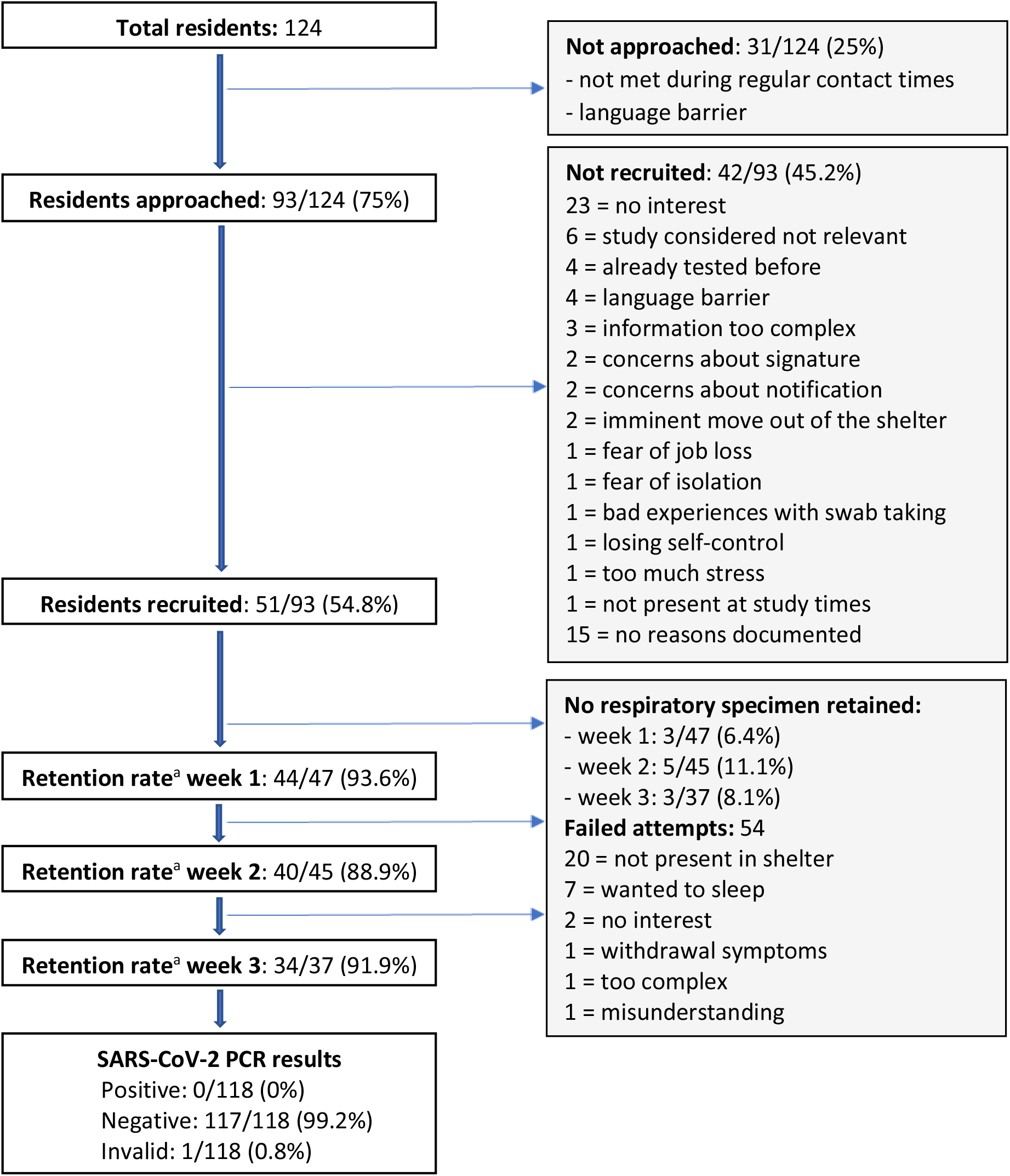
Study flow diagram with reasons for non-recruiting and non-retaining of residents for a weekly respiratory specimen. ^a^ retention rate: number of residents monitored with analysis of a respiratory specimens per week compared with the number of recruited residents that were still living in the shelter during that week

Baseline characteristics of the potential participants and the informed consent process are displayed in table 2. Seventy-four individuals (79.6%) were male, with a male:female ratio of 4.9 : 1; median age was 47 years (IQR 34─54), with a range from 21 to 86 years. Information of participants took place mainly in the evening (94.6%). One third (33.3%) received the information in a group. Median estimated duration of the information was 10 minutes (IQR 5─10). 55.9% of the potential participants received information in other languages than German, of which all were available within the co-researcher team: Russian (18.3%), English (17.2%), Polish (12.9%), Romanian (6.5%) and French (1.1%).

**Table 2.** Baseline characteristics of residents approached for participation and of the informed consent process

|  |  | Residents approached (n=93) |
| --- | --- | --- |
| **Sex** | Female | 15 (16.1%) |
|  | Male | 74 (79.6%) |
| **Age, median, years (IQR)** |  | 47 (34-54) |
| **Daytime of information** | Morning | 2 (2.2%) |
|  | Noon | 0 |
|  | Evening | 88 (94.6%) |
| **Individual information** |  | 59 (63.4%) |
| **Group information** |  | 31 (33.3%) |
| **Language mediation** | Yes | 53 (57%) |
|  | No | 37 (39.8%) |
| **Language of consultation** | German | 37 (39.8%) |
|  | Russian | 17 (18.3%) |
|  | English | 16 (17.2%) |
|  | Polish | 12 (12.9%) |
|  | Romanian | 6 (6.5%) |
|  | French | 1 (1.1%) |
| **Duration, median, minutes (IQR)** |  | 10 (5-10) |
| **Questions of participants** | 3 = general procedure of the study<br>2 = communication of results<br>2 = times of specimen collection<br>1 = potential costs<br>1 = aim of study |  |
| **Concerns of participants** | 2 = use of personal data<br>1 = giving signature |  |
| **Difficulties perceived by co-researcher team** | 9 = difficulties in communication<br>3 = uncertainty<br>2 = moving out soon<br>2 = lack of interest |  |
| **Consent to participation** | Yes | 51 (54.8%) |
|  | No | 42 (45.2%); 5 later withdrawn |

Data are n (%); age and duration with median (interquartile range). Missing data: sex (n=4), daytime (n=3), individual/group information (n=3), language spoken (n=4).

Frequently asked questions by the residents addressed the general procedure (n=3), and aim of the study (n=1), communication of results (n=2) and times of specimen collection (n=2). Some had concerns about the use of personal data (n=2) and signing the consent form (n=1). Difficulties perceived by the co-researcher team were communication (n=9), uncertainty of the resident (n=3), move out of the shelter (n=2), and lack of interest (n=2). A written informed consent could be obtained from 51 out of 93 (54.8%) of the potential participants.

Consenting participants were comparable to non-consenting participants by age and spoken language. There were more women among the non-consenters (n=9, [21,4%]) than among the consenters (n=6, [11.8%]). The reasons for refusing participation, as perceived by the study team, are shown in Fig. 1.

### Retention and acceptability

During the first week, a respiratory specimen (saliva) from 44 out of 47 (93.6%) residents who had consented and were living in the shelter during that week could be retained for SARS-CoV-2 testing by RT-PCR. During the second and third week, a respiratory specimen (self-collected swab of the tongue, buccal mucosa, and anterior nares) from 40 out of 45 (88.9%) and from 34 out of 37 (91.9%) residents, respectively, could be retained for testing.

In many cases, repeated attempts to collect the specimen were necessary. Fifty-four failed attempts of specimen collection were documented by the co-researcher team, mainly because the resident was not present in the shelter (n=20) or because the collecting time of the samples in the morning was inconvenient (n=7). Furthermore, in some cases, a specimen collection was not possible due to a lack of interest (n=2), withdrawal symptoms (n=1), misunderstanding of study procedure (n=1) or the complexity of sampling (n=1) despite assistance (Figure 1). Dry mouth and, therefore, long duration of sampling was mentioned to be difficult in 5 cases as well as the use of a straw for saliva collection (n=2). According to the focus group, difficulties during the specimen collection were underreported in the evaluation forms. Early morning collection of samples was perceived as a burden for both residents and staff. A flexible collection of samples during the day would have been preferred.

For the self-collected swab of the tongue, buccal mucosa and anterior nares, 24 out of 52 (46.2%) residents refused to place the swab in the nares after having been in the oral cavity for hygienic reasons or perceived as uncomfortable. The co-researcher team pointed out that lack of fine motoric skills made the self-collection difficult in some cases.

### Specimens and results of RT-PCR analysis

Sample self-collection was guided by an instruction leaflet provided to the participants [see Additional files 1, 2]. The targeted volume of saliva was 2 ml per specimen and marked on the collection tube. The median volume collected was 1.7 ml (IQR 0.9-2), thereof 7 specimens with a volume of less than 0.5 ml. The visual inspection of the specimen revealed the following abnormalities: empty tube (n=1), tube not adequately closed (n=1), obvious external contamination with saliva (n=1), specimen consistent and viscous (n=6). The visual inspection of the self-collected swab from tongue, buccal mucosa, and anterior nares revealed the following abnormalities: tube not adequately closed (n=1) and media in tube incomplete (n=1). The highly viscous nature of saliva samples (at least at the time when samples arrived in the laboratory) made it necessary to pre-treat samples before routine molecular diagnostic testing. Thus, this sample type is more prone to invalid test results than the eSwabs used in the second and third week. This is especially true for sample processing in fully automized high throughput testing systems like Roche’s cobas 6800/8800.

During the study period, no resident was tested positive for SARS-CoV-2 by RT-PCR. 117/118 (99.2%) specimens were tested negative. One analysis was considered invalid as the sample tube appeared to be empty.

### Workload

During the 3-weeks of the pilot study, a total of about 274 person-hours were invested by the co-researcher team for training and meetings (39.1%), obtaining informed consent (32.8%), weekly sample collection including preparation of material (17.5%), project coordination (7.8%) and data maintenance (2.8%). The workload was distributed between 2 project coordinators and 20 team members. The working hours of the supervision team were not included in the analysis. In the focus group and final evaluation questionnaire it was emphasized that the workload was high and difficult to manage together with other routine activities.

### Feasibility and critical appraisal

Seventeen out of 22 co-researcher team members participated in a final focus group discussion. Overall, the implementation of the monitoring was perceived to be valuable and a good experience, but work-intensive. Only 1 out of 17 team members stated that monitoring would not be possible during the winter season in the emergency night shelters. In the critical review of the results, the team reported that barriers for recruitment and retention were underestimated due to incomplete documentation in the evaluation forms. Non-availability of a common language, particularly Bulgarian, Lithuanian, Czech, and Vietnamese, was the main barrier to participation. Provision of written negative test results could have increased participation among the residents. Also, the presence of people in the team who were familiar to the residents was crucial and increased trust and willingness to participate.

Twelve team members participated in the final evaluation with a structured questionnaire. Eleven out of 12 preferred the self-collected swab for the following reasons: more flexible times of collection (n=6), more hygienic (n=5), less complicated (n=5), and faster to collect (n=4). Visual instruction leaflets for sampling [see Additional files 1, 2] were perceived as suitable by 7 team members, whereas on-site demonstration and direct assistance during the collection was perceived to be helpful or necessary by 10 team members. The use of video or audio formats was suggested (n=6) as optimization of information and instruction for sample collection.

Table 3 provides selected themes with illustrative quotes that were emphasized by the co-researchers in the focus group, online platform, or final evaluation questionnaire. In terms of acceptability by the residents, allowing flexible times of sample collection or of providing information, as well as adapting the written information in a more precise, adapted language was suggested.

**Table 3.** Selected quotes emphasized by the co-researchers in the focus group, online platform, or final questionnaire

| Theme | Quotes |
| --- | --- |
| Interaction with residents | *The project team should include people who have already built up trust to the homeless community* |
|  | *Be prepared for multiple languages and guarantee barrier-free communication* |
|  | *Use audio- and video formats for provision of information* |
|  | *Informed consent and sample collection may be influenced by intoxication of residents* |
|  | *It's a nice way to talk to people you didn't know before. Many were happy about the conversation* |
| Willingness for participation | *During the cold season, homeless people might have other priorities, consider needs and daily routine of the residents in the planning* |
| Specimen collection | *Patience and understanding for repeated instructions are needed* |
|  | *With more flexible times of sample collection, we could have reached more residents* |
|  | *Without assistance, the collection would have not been possible for several residents* |
| Staff and workload | *Good and continuous communication among the health workers, social workers and language mediators, as well as supervision was essential* |
|  | *Additional staff is needed due to high workload of monitoring activities* |
|  | *Benefits for residents and staff should be identified and emphasised. The monitoring gave me a feeling of security in the shelter.* |
| Ethical considerations | *Information on data use and consequences of a positive test result should be transparent* |
|  | *Accept a “no”, take people as they are* |

## Discussion

### Summary of main results

Fifty-one out of 93 (54.8%) residents were recruited for this study. High retention rates (88.9% – 93.6%) of a weekly respiratory specimen could be reached during the 3 weeks, however, in many cases repeated attempts to collect the specimen were necessary. A self-collected swab of the tongue, buccal mucosa and anterior nares was considered easier and more hygienic to collect than a saliva specimen. Several of the saliva samples showed a reduced volume and high viscosity making them less suitable for standardized molecular diagnostic testing. On-site demonstration and assistance were frequently necessary with both saliva and swab in order to obtain an adequate specimen. A considerable number of residents (n=25, [46.2%]) refused to place the swab in the nares after having placed it in the oral cavity due to hygienic reasons or because it was perceived as uncomfortable. All specimens tested were negative for SARS-CoV-2 by RT-PCR during a period of low community transmission in Berlin in July 2020 [30].

There were more women among the non-consenters (n=9, [21,4%]) than among the consenters (n=6, [11.8%]). Two-thirds of conversations were conducted with language mediation, mainly in Eastern European languages. Language barriers were one of the main reasons for non-recruitment and difficulties in the recruitment process. Barriers for participation were lack of interest to participate, perception that study information was too complex, concerns about the use of personal data, and providing a signature. Flexibility of sample collection schedules, the use of video and audio materials, and concise written information were the main recommendations of the co-researchers for future implementation. Sufficient human resources were considered essential for the successful implementation of a monitoring concept, which also allows the individual needs of the residents to be considered.

### Strengths and limitations

An understanding of the challenges and issues related to recruitment and retention – especially in a so called hard-to-reach population – is important and can help policy makers to foresee strategies to overcome these issues. In Berlin, this may be relevant for the reopening of > 40 emergency overnight shelters in the winter season 2020/21 [28].

The residents of the 24/7 shelter were not representative of all people experiencing homelessness in Berlin. People in highly precarious situations (e.g., with psychiatric disorders) were unlikely fully represented, due to the fact that individuals had to register at the shelter and fulfill certain requirements during their stay. The need of a signature in the study consent form may have discouraged some individuals from participating. The high retention rates may not be fully generalizable, as the co-researchers put huge efforts in conducting the study, demonstrated by the repeated attempts to collect the specimens. The analysis of recruitment and retention barriers may be biased, as it relied on observations made by the co-researchers. Lack of interest to participate or indifference to the topic might have different reasons such as lack of adequate information, or other basic priorities. The interval of a weekly respiratory specimen was a pragmatic decision, as it remains unclear which intervals would most efficiently prevent chains of infection. People experiencing homelessness were not involved in the planning, implementation, or the evaluation of this study.

### Implications for future monitoring concepts

In the USA, COVID-19 outbreaks have been observed in homeless shelters with high proportions of infected residents and staff, including a high number of asymptomatic individuals [7, 11, 12]. Testing to diagnose COVID-19 is only one component of a required comprehensive infection prevention strategy including for example promoting behaviors that reduce transmission or specific quarantine housing. There are different testing strategies that can be applied in homeless shelters. Firstly, testing of residents with symptoms consistent with COVID-19; symptom screening may help to identify those individuals. Secondly, testing asymptomatic residents with exposure to SARS-CoV-2. In regard to challenges in tracing close contacts within homeless shelters, broader testing of residents and staff, e.g., facility-wide testing, can be considered [15]. Thirdly, testing asymptomatic residents without known exposure may allow for early identification of COVID-19 cases and outbreaks. If there is moderate or substantial transmission in the community, initial and regular shelter-wide (universal) testing may be considered for transmission control [12, 15]. The CDC does not recommend entry testing for homeless service sites, as the additional benefit to other implemented preventive measures is unknown [15]. Risk mitigation strategies of COVID-19 outbreaks, including universal testing in high-risk facilities, will require flexible adapted approaches that have to consider the level of the community transmission, the available resources, and the local recommendations. Microsimulation models may help to argue about the impact, costs, and cost-effectiveness of universal testing in homeless shelters according to various scenarios [31].

The workload for the co-researchers was high and difficult to manage together with other routine activities. Besides human resources, limited laboratory capacity for RT-PCR analysis may be a major barrier for the realization of monitoring concepts, especially if mass individual testing would be envisaged. Testing multiple samples in one approach (pooling) to screen asymptomatic people is an important strategy to consider – even if associated with challenges – when testing capacity is low and laboratory infrastructure overwhelmed [32-36]. It is also a more socially responsible strategy in regard to limited testing capacity globally [37].

Testing strategies should be implemented in a way that protects privacy and confidentiality [15]. The vulnerability of the homeless population in terms of discrimination, social exclusion, residency status and resulting dependencies has to be considered in program planning. Especially in obtaining informed consent, all relevant information should be provided in an appropriate and understandable way respecting the autonomy of the shelter resident.

## Conclusion

This study demonstrated that with appropriate efforts voluntary, regular universal testing for SARS-CoC-2 is feasible in homeless shelters. In our opinion, there are key points for successful implementation. The value of monitoring COVID-19 has to be emphasized to promote understanding and acceptability of testing, as residents may have other, more pending basic needs, especially during the winter season. Language barriers must be specially addressed, including the use of digital formats. A less invasive sampling method will result in higher compliance for regular swab testing. Self-testing with assistance, like undertaken in this study, also requires significantly less resources of qualified staff and personal protective equipment. In the current situation of a pandemic, monitoring concepts will have to accept a possible lower sensitivity of the applied methods in order to be feasible and to allow screening of individuals without symptoms on a broader scale who might spread SARS-CoV-2. Universal testing of high-risk facilities should be considered according to the level of community transmission and the available resources. Finally, participatory approaches should be sought with monitoring strategies that consider homeless people’s needs and life situation.

## Data Availability

The datasets generated and/or analyzed during the current study are not publicly available due containing information that could compromise the privacy/consent of the shelter residents but are available from the corresponding author on reasonable request.

## Abbreviations

COVID-19: Coronavirus Disease 2019
SARS-CoV-2: Severe Acute Respiratory Syndrome Coronavirus Type 2
RT-PCR: Reverse Transcription-Polymerase Chain Reaction
IQR: Interquartile Range
CDC: Centers for Disease Control and Prevention

## Declarations

### Ethics approval and consent to participate

This study was approved by the ethics committee of the Charité – Universitätsmedizin (No.: EA4/141/20). Written informed consent to participate in the study was obtained.

### Consent for publication

Not applicable.

### Availability of data and materials

The datasets generated and/or analyzed during the current study are not publicly available due containing information that could compromise the shelter residents’ privacy/consent but are available from the corresponding author on reasonable request.

### Authors’ contributions

A.K.L. and N.S. designed and supervised the study, led the data analysis and the writing of the manuscript. Both contributed equally. All authors contributed to the conduct of the study and the writing of the manuscript.

## Acknowledgements

Derrick Akechu Wouba, Gabriela Aldama, Oskar Herbst, Pearl von Herder, Sophie Hilt, Anna Behnke, Wojciech Greh, Leon Hoffmann, Dominika Jurasik, Miriam Luchterhand, Jonas Kalmbach, Valeska Steinert, Sophie Rothe, Uldis Stukmanis, Franek Machowski, Olga Nikolai, Heike Rössig.

## Supplementary Information (in the final publication)

**Additional file 1:** Visual instruction leaflet for self-collection of saliva

**Additional file 2:** Visual instruction leaflet for a self-collected swab of tongue, buccal mucosa and anterior nares

**Additional file 3:** Description of the coding tree for the qualitative analysis

**Additional file 4:** Responses to the consolidated criteria for reporting qualitative research (COREQ)

